# A haemagglutination test for rapid detection of antibodies to SARS-CoV-2

**DOI:** 10.1101/2020.10.02.20205831

**Authors:** Alain Townsend, Pramila Rijal, Julie Xiao, Tiong Kit Tan, Kuan-Ying A Huang, Lisa Schimanski, Jiangdong Huo, Nimesh Gupta, Rolle Rahikainen, Philippa C Matthews, Derrick Crook, Sarah Hoosdally, Teresa Street, Justine Rudkin, Nicole Stoesser, Fredrik Karpe, Matthew Neville, Rutger Ploeg, Marta Oliveira, David J Roberts, Abigail A Lamikanra, Hoi Pat Tsang, Abbie Bown, Richard Vipond, Alexander J Mentzer, Julian C Knight, Andrew Kwok, Gavin Screaton, Juthathip Mongkolsapaya, Wanwisa Dejnirattisai, Piyada Supasa, Paul Klenerman, Christina Dold, Kenneth Baillie, Shona C Moore, Peter JM Openshaw, Malcolm G Semple, Lance CW Turtle, Mark Ainsworth, Alice Allcock, Sally Beer, Sagida Bibi, Elizabeth Clutterbuck, Alexis Espinosa, Maria Mendoza, Dominique Georgiou, Teresa Lockett, Jose Martinez, Elena Perez, Veronica Sanchez, Giuseppe Scozzafava, Alberto Sobrinodiaz, Hannah Thraves, Etienne Joly

**Author notes:** Corresponding Authors Correspondence to Alain Townsend ( k) and Etienne Joly.

## Abstract

Serological detection of antibodies to SARS-CoV-2 is essential for establishing rates of seroconversion in populations, detection of seroconversion after vaccination, and for seeking evidence for a level of antibody that may be protective against COVID-19 disease. Several high-performance commercial tests have been described, but these require centralised laboratory facilities that are comparatively expensive, and therefore not available universally. Red cell agglutination tests have a long history in blood typing, and general serology through linkage of reporter molecules to the red cell surface. They do not require special equipment, are read by eye, have short development times, low cost and can be applied as a Point of Care Test (POCT). We describe a red cell agglutination test for the detection of antibodies to the SARS-CoV-2 receptor binding domain (RBD). We show that the Haemagglutination Test (“HAT”) has a sensitivity of 90% and specificity of 99% for detection of antibodies after a PCR diagnosed infection. The HAT can be titrated, detects rising titres in the first five days of hospital admission, correlates well with a commercial test that detects antibodies to the RBD, and can be applied as a point of care test. The developing reagent is composed of a previously described nanobody to a conserved glycophorin A epitope on red cells, linked to the RBD from SARS-CoV-2. It can be lyophilised for ease of shipping. We have scaled up production of this reagent to one gram, which is sufficient for ten million tests, at a cost of ∼0.27 UK pence per test well. Aliquots of this reagent are ready to be supplied to qualified groups anywhere in the world that need to detect antibodies to SARS-CoV-2, but do not have the facilities for high throughput commercial tests.

## INTRODUCTION

Red cell agglutination tests have a distinguished history. Since Landsteiner’s classic observations in 1901 (Landsteiner, 1961) (English translation), they have been used for the determination of blood groups (Schwarz & Dorner, 2003) detection of influenza viruses (Hirst, 1941) and in a wide variety of applications championed by Prof. Robin Coombs for the detection of specific antibodies or antigens (Coombs, Mourant, & Race, 1945) (reviewed by (Pamphilon & Scott, 2007)). They have the great advantage of being simple, inexpensive, can be read by eye, and do not require sophisticated technology for their application. In the recent era the linkage of an antigen to the red cell surface has become easier with the possibility of fusing a protein antigen sequence with that of a single domain antibody or nanobody specific for a molecule on the red cell surface (discussed in (Habib et al., 2013)).

We have applied this concept to provide a simple Haemagglutination Test (“HAT”) for the detection of antibodies to the Receptor Binding Domain (RBD) of the SARS-CoV-2 spike protein. The RBD is a motile subdomain at the tip of the SARS-CoV-2 spike protein that is responsible for binding the virus to its ACE2 receptor. The RBD of betacoronaviruses folds independently of the rest of the spike protein (Lan et al., 2020; Li, Li, Farzan, & Harrison, 2005; Wang et al., 2020; Yan et al., 2020). This useful property provides an Achilles’ heel for the virus and allows many potential applications in vaccine design (Dai et al., 2020; Mulligan et al., 2020; Tan et al., 2020; Walls et al., 2020; Yang et al., 2020; Zha et al., 2020), and serology (Amanat et al., 2020; Piccoli et al., 2020; The National SARS-CoV-2 Serology Assay Evaluation Group, 2020), see also www.gov.uk/government/publications/COVID-19-laboratory-evaluations-of-serological-assays). The majority of neutralising antibodies bind to the RBD (Barnes et al., 2020; Piccoli et al., 2020), and the level of antibody to the RBD detected in ELISA correlates with that of neutralising antibodies (Amanat et al., 2020; Piccoli et al., 2020; Robbiani et al., 2020). We reasoned therefore that a widely applicable and inexpensive test for antibodies to the RBD would be useful for research in settings where high throughput assays were not available.

In order to link the SARS-CoV-2 RBD to red cells we selected the single domain antibody (nanobody) IH4 (Habib et al., 2013), specific for a conserved epitope on glycophorin A. Glycophorin A is expressed at up to 10^6^ copies per red cell. The IH4 nanobody has previously been linked to HIV p24 to provide a monomeric reagent that bound p24 to the red cell surface. Antibodies to p24 present in serum cross-linked the p24 and agglutinated the red cells (Habib et al., 2013). We have adapted this approach to detection of antibodies to SARS-CoV-2 by linking the RBD of the SARS-CoV-2 spike protein to IH4 via a short (GSG)2 linker to produce the fusion protein IH4-RBD-6H (Figure 1). Since we embarked on this project, another group has described preliminary results with an approach similar to ours, but using a fusion of the RBD to an ScFV against the H antigen to coat red blood cells with the SARS-2 RBD (Kruse et al., 2020).

**Figure 1.**
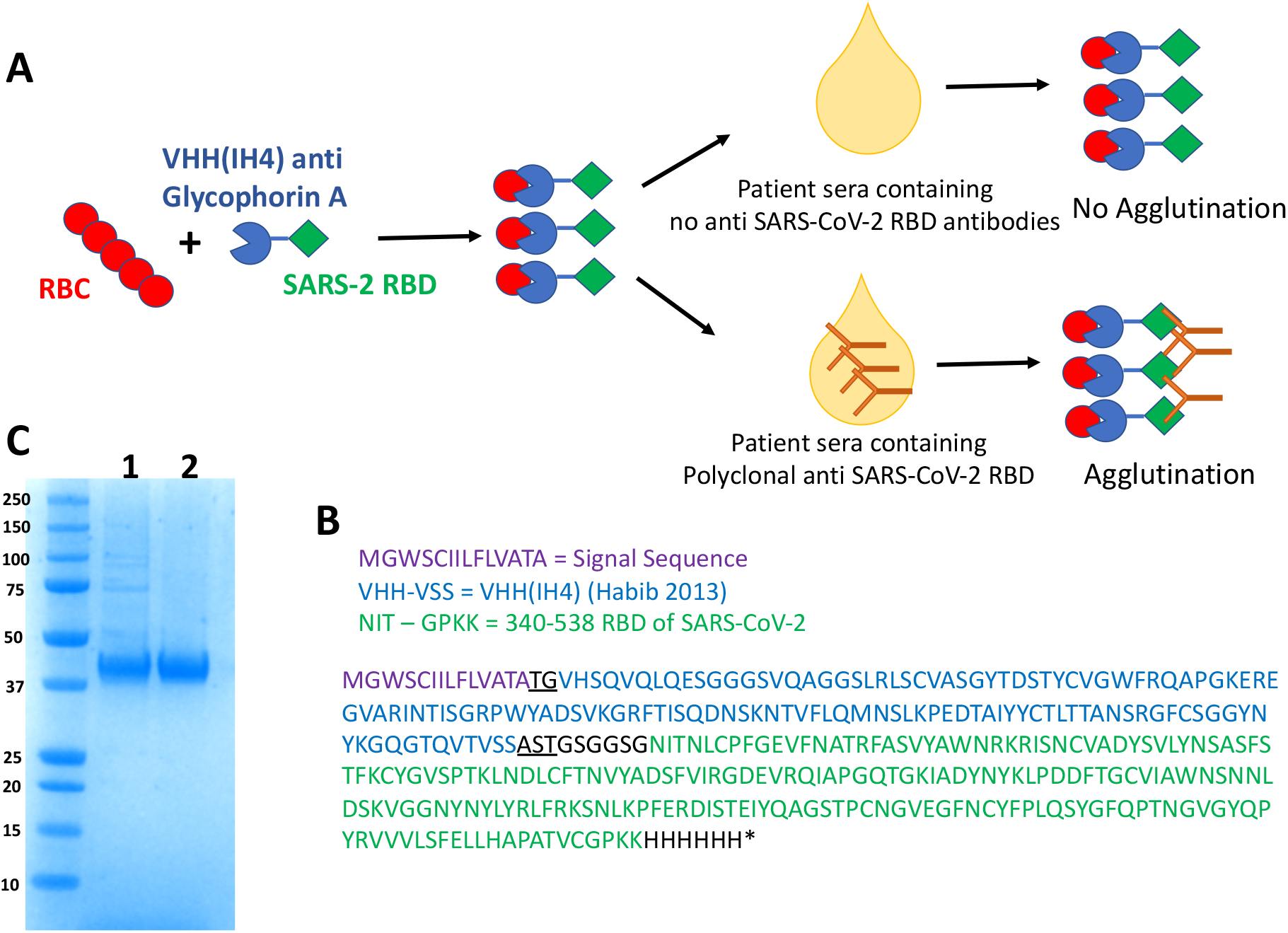
Haemagglutination Test (HAT) for detection of antibodies to SARS-CoV-2 Receptor Binding Domain. A) Concept of the HAT B) Sequence of VHH(IH4)-RBD fusion protein. Residues underlined are encoded by cloning sites AgeI (TG) and SalI (AST). The codon optimised cDNA sequence is shown in supplementary Information C) SDS-PAGE gel of purified VHH(IH4)-RBD proteins. Three micrograms of protein were run on 4-12% Bolt Bis-Tris under reducing conditions. 1: IH4-RBD produced in house in Expi293F cells, 2: IH4-RBD produced by Absolute Antibody, Oxford in HEK293 cells.

## RESULTS

### Production of the IH4-RBD Reagent

The IH4-RBD sequence (Figure 1B) was codon optimised and expressed in Expi293F cells in a standard expression vector (available on request). One advantage of this mode of production compared to bacterially produced protein as used by Habib et al, is that the reagent will carry the glycosylation moieties found in humans, which may play a role in the antigenicity of the RBD (Pinto et al., 2020). The protein (with a 6xHis tag at the C-terminus for purification) was purified by Ni-NTA chromatography which yielded ∼160 mg/L. We later had one gram of the protein synthesised commercially by Absolute Antibody, Oxford. The IH4-RBD protein ran as single band at ∼40 kDa on SDS PAGE (Figure 1C).

### Establishment of the Haemagglutination Test (HAT) with monoclonal antibodies to the RBD

One purpose envisaged for the HAT is for use as an inexpensive Point of Care Test for detection of antibodies in capillary blood samples obtained by a “finger-prick”. We therefore wished to employ human red cells as indicators without the need for cell separation or washes, to mimic this setting. The use of V-bottom microtiter plates to perform simplified hemagglutination tests was first described over 50 years ago (Wegmann & Smithies, 1966). In preliminary tests, we observed that 50 μL of whole blood (K2EDTA sample) diluted 1:40 in phosphate-buffered saline (PBS), placed in V-bottomed wells of a standard 96-well plate, settled in one hour to form a button of red cells at the bottom of the well. The normal haematocrit of blood is ∼40% vol/vol, so this dilution provides ∼1% red cells. If the plate was then tilted, the red cell button flowed to form a “teardrop” in ∼30 seconds (for example Figure 2A Row 8).

**Figure 2.**
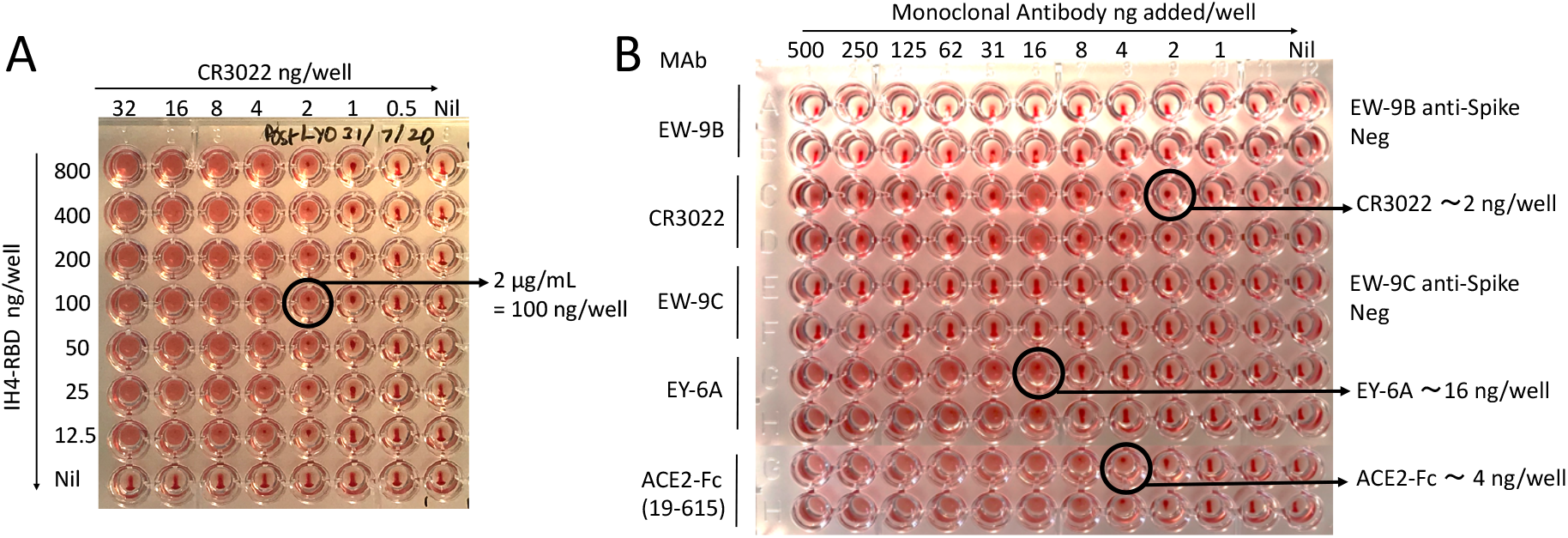
Haemagglutination with human monoclonal antibodies or nanobodies to the SARS-CoV-2 RBD. A) Titration of IH4-RBD and monoclonal Antibody CR3022 to RBD. Doubling dilutions of CR3022 and IH4-RBD were prepared in separate plates. 50 μL red cells (O-ve whole blood diluted 1:40 in PBS) were added to the CR3022 plate, followed by transfer of 50 μL titrated IH4-RBD. From this titration, 100 ng/well of IH4-RBD was chosen for detection. B) Detection of other anti-RBD monoclonal antibodies and ACE2-Fc. Monoclonal antibodies were prepared in doubling dilutions in 50 μL PBS from left to right, 50 μL of 1:40 O-ve red cells were added, followed by 50 μL of IH4-RBD (2 μg/mL in PBS). The end point was defined as the last dilution without tear drop formation on tilting the plate for ∼ 30 s. The binding sites for CR3022, EY6A and ACE2 on RBD have been defined (Huo, Zhao, et al., 2020; Lan et al., 2020; Yan et al., 2020; Zhou et al., 2020). EW-9B and EW-9C are monoclonal antibodies against non-RBD epitopes on the spike protein (Huang et al., 2020). ACE2-Fc has been described (Huang et al., 2020).

If serum or plasma samples are to be tested, a standard collection of 10 mL of Type O Rh-negative (O-ve) blood into a K2EDTA tube will thus provide sufficient red cells for 8,000 test wells.

A well characterised monoclonal antibody to the RBD, CR3022 (ter Meulen et al., 2006) added to the red cells at between 0.5-32 ng/well in 50 μL, did not agglutinate the cells on its own (Figure 2A Row 8). The addition of the IH4-RBD reagent at between 12.5-800 ng/well (in 50 μL PBS) induced a concentration dependent agglutination of the red cells, detected by the formation of a visible mat or plug of agglutinated cells, and the loss of teardrop formation on tilting the plate (Figure 2A). From repeated trials of this experiment we established that a standard addition of 100 ng/well of the IH4-RBD developer (50 μL of a stock solution of 2 μg/mL in PBS) induced agglutination of 50 μL of 1:40 human red cells in the presence of as little as 2 ng/well of the CR3022 monoclonal antibody. The standardised protocol used for the subsequent tests were thus performed in 100 or 150 μL final volume, containing 100 ng of the IH4-RBD developer, and 50 μL 1:40 whole blood (∼1% v/v red cells ∼ 0.5 μL packed red cells per reaction). After 60 minutes incubation at room temperature, we routinely photographed the plates after the 30 s tilt for examination and reading.

The requirement for 100 ng of the IH4-RBD developer per test well means that the gram of IH4-RBD protein we have had synthesised is sufficient for 10 million test wells at a cost of approximately 0.27 UK pence per test.

Having established a standard addition of 100 ng/well of the IH4-RBD reagent, we screened a set of twelve human monoclonal antibodies, two divalent nanobodies, and divalent ACE2-Fc, that are known to bind to the RBD (Huang et al., 2020; Huo, Le Bas, et al., 2020; Pinto et al., 2020; Wrapp et al., 2020; Zhou et al., 2020). These reagents bind to at least three independent sites on the RBD, and some are strongly neutralising and capable of profound ACE2 blockade (Huang et al., 2020; Huo, Le Bas, et al., 2020). Twelve of the 15 divalent molecules agglutinated red cells and titrated in the HAT to an end point between 2-125 ng/well, after addition of 100 ng IH4-RBD (Figure 2B and Supplementary Table 1). Two monoclonal antibodies, FD-5D and EZ-7A (Huang et al., 2020) and one divalent nanobody VHH72-Fc (Wrapp et al., 2020), failed to agglutinate red cells in the presence of the IH4-RBD reagent. However, if a monoclonal antibody to human IgG was added to the reaction (50 μL of mouse anti-human IgG, Sigma Clone GG5 1:100), these molecules specifically agglutinated the red cells (Supplementary Figure 1A, B). This result, analogous to the “indirect” Coombs Test (Coombs et al., 1945; Pamphilon & Scott, 2007), suggested that these three molecules had bound to the RBD associated with the red cells but failed to crosslink to RBDs on neighbouring red cells. However, these could be crosslinked by the anti IgG reagent. Monoclonal antibodies to other regions of the spike protein (EW-9B, EW-9C and FJ-1C) failed to agglutinate red cells (Figure 2B, Supplementary Table 1). Finally we looked at the effect of a divalent ACE2-Fc molecule constructed by fusing the peptidase domain of ACE2 (amino acids 19-615) to the hinge and Fc region of human IgG1 (described in (Huo, Le Bas, et al., 2020). ACE2-Fc agglutinated red cells strongly in the presence of 100 ng/well of the IH4-RBD developer, titrating to ∼ 4 ng/well (Figure 2B rows 7,8).

In summary, these results showed that all of the known epitopes bound by characterised monoclonal antibodies were displayed by the IH4-RBD reagent, as well as the ACE2 binding site, and could mediate agglutination by specific antibodies, divalent nanobodies, or ACE2-Fc.

### Agglutination by plasma from donors convalescing from COVID-19

These experiments established the conditions for detection of haemagglutination by monoclonal antibodies to the RBD, in particular the optimum concentration of IH4-RBD of 100 ng/well. We then proceeded to look for haemagglutination by characterised plasma from COVID-19 convalescent donors. In the first trial we tested eighteen plasma samples from patients with mostly mild illness, that had been characterised with a quantitative ELISA to detect antibodies to the RBD (Peng et al., 2020). For these experiments we used fresh O-ve blood (K2EDTA sample) diluted to 1:40 as a source of red cells to avoid agglutination by natural agglutinins in the plasma. Plasmas were titrated by doubling dilution from 1:20 in 50 μL, then 50 μL of 1:40 O-ve red cells were added, followed by addition of 100 ng of the IH4-RBD in 50 μL PBS. After one-hour incubation, plates were tilted for ∼30 seconds, photographed and read. The titre of agglutination was assessed by complete loss of teardrop formation by the red cells, any formation of a teardrop was regarded as negative. Figure 3A shows that the HAT titre matched the RBD ELISA results. Four samples were scored as negative in both assays. The remaining results showed that in general the HAT titre increased with the ELISA end point titre. One sample gave a positive titre of 1:320 in the HAT but was negative in ELISA (indicated with an arrow). We investigated this sample with a developer composed of the IH4 nanobody without the RBD component, which revealed that agglutination was RBD dependent (not shown). This sample was also positive at 1:1123 in an ELISA for full length spike protein (not shown), which suggests that the antibodies contained in this serum recognised epitope(s) present on the RBD exposed in the HAT, but not on the RBD in the RBD-ELISA reference test (Peng et al., 2020). The highest titre detected in these samples by the HAT was 1:1280 (Figure 3B).

**Figure 3.**
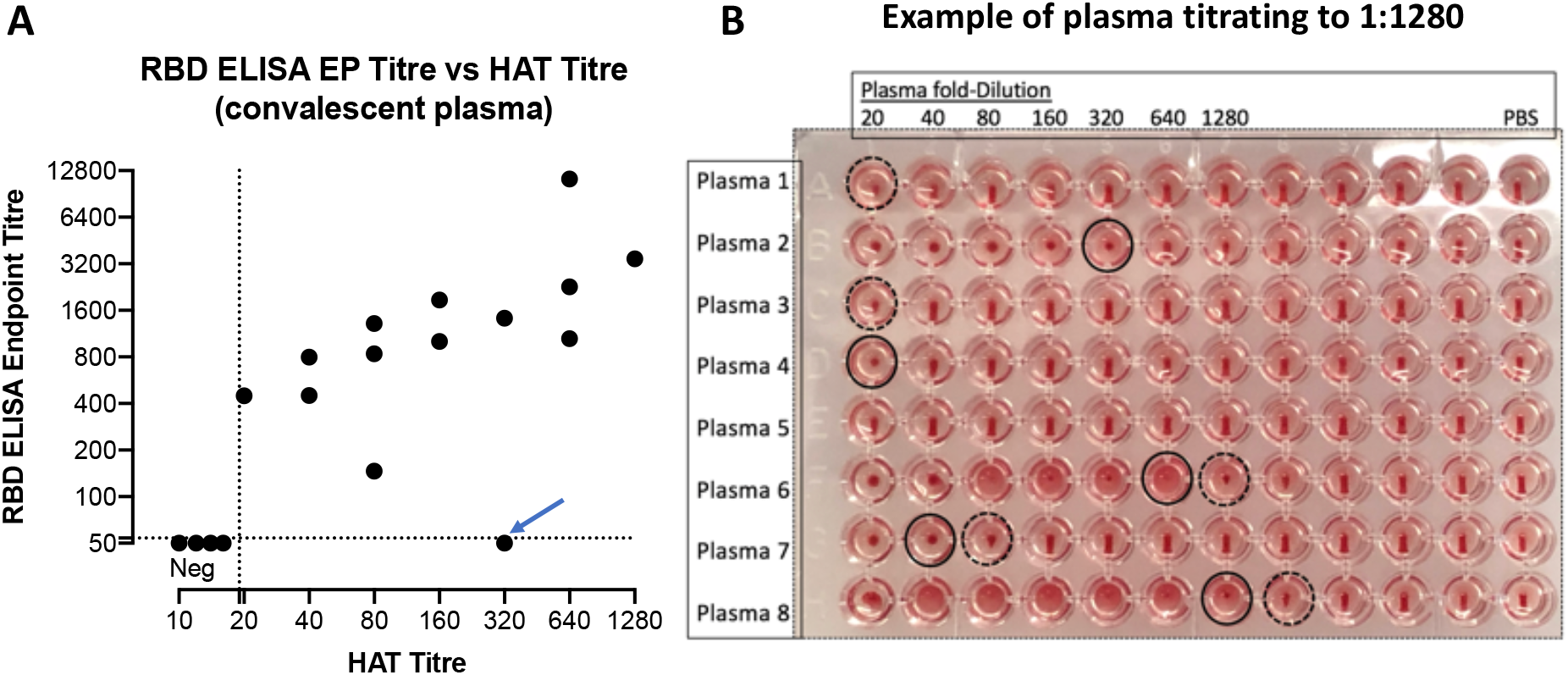
Titration of stored plasma in the agglutination assay. A) Eighteen plasma samples from mild cases were compared for titration in the HAT with 1:40 O-ve whole blood from a seronegative donor, and endpoint titre in an RBD ELISA (Peng et al., 2020)). Four samples were negative in both assays. The data point marked with an arrow on the graph (plasma 2 on the plate, Fig 3B) was checked with a reagent composed of IH4 without RBD and shown to be dependent on antibodies to the RBD. This sample did score positive for antibodies to full length spike in an ELISA (EPT 1:1123). B) An example of titration: positive agglutination endpoints (loss of teardrop) are marked with a black solid-line circle, partial teardrops are marked with a dotted-line circle.

These preliminary results showed that the HAT could detect antibodies to the RBD in plasma samples from convalescent patients in a similar manner to an ELISA test, but were not sufficient to establish the sensitivity and specificity of the HAT.

### Sensitivity and specificity of the Haemagglutination Test

To formally assess the sensitivity and specificity of the HAT we collected a set of 98 “positive” plasma samples from donors diagnosed with COVID-19 by RT-PCR at least 28 days prior to sample collection (NHS Blood and Transplant), and 199 “negative” serum samples from healthy donors from the pre-COVID-19 era (Oxford Biobank). The samples were randomised before plating. The test wells were arranged in duplicate to contain serum/plasma at 1:40 dilution, and 1:40 O-ve red cells in 50 μL. 100 ng of IH4-RBD in 50 μL PBS was added to one well of the pair, 50 μL of PBS to the other (as a negative control). The negative control is important because in rare cases, particularly in donors who may have received blood transfusions, the sample may contain antibodies to non-ABO or Rhesus D antigens. After development, the plates were photographed, and read by two independent masked observers. Complete loss of teardrop was scored as positive, any flow in the teardrop as negative. These rules were established before setting up the tests. Disagreements (7% overall) were resolved by accepting the weaker interpretation – i.e. if one observer scored positive but the other negative, the well was scored as negative. Having completed the scoring the columns of samples were re-randomised and the test and scoring repeated.

Examples of test wells and scoring are shown in Figure 4. The red cells in the negative control (PBS) wells formed a clear teardrop. Red cells in positive wells (indicated with a solid ring) settled either into a mat or a button that failed to form any teardrop on tilting for 30 seconds. Occasional wells (15 of 297) formed a “partial” teardrop (shown by a dashed ring). These were scored as negative by prior agreement.

**Figure 4.**
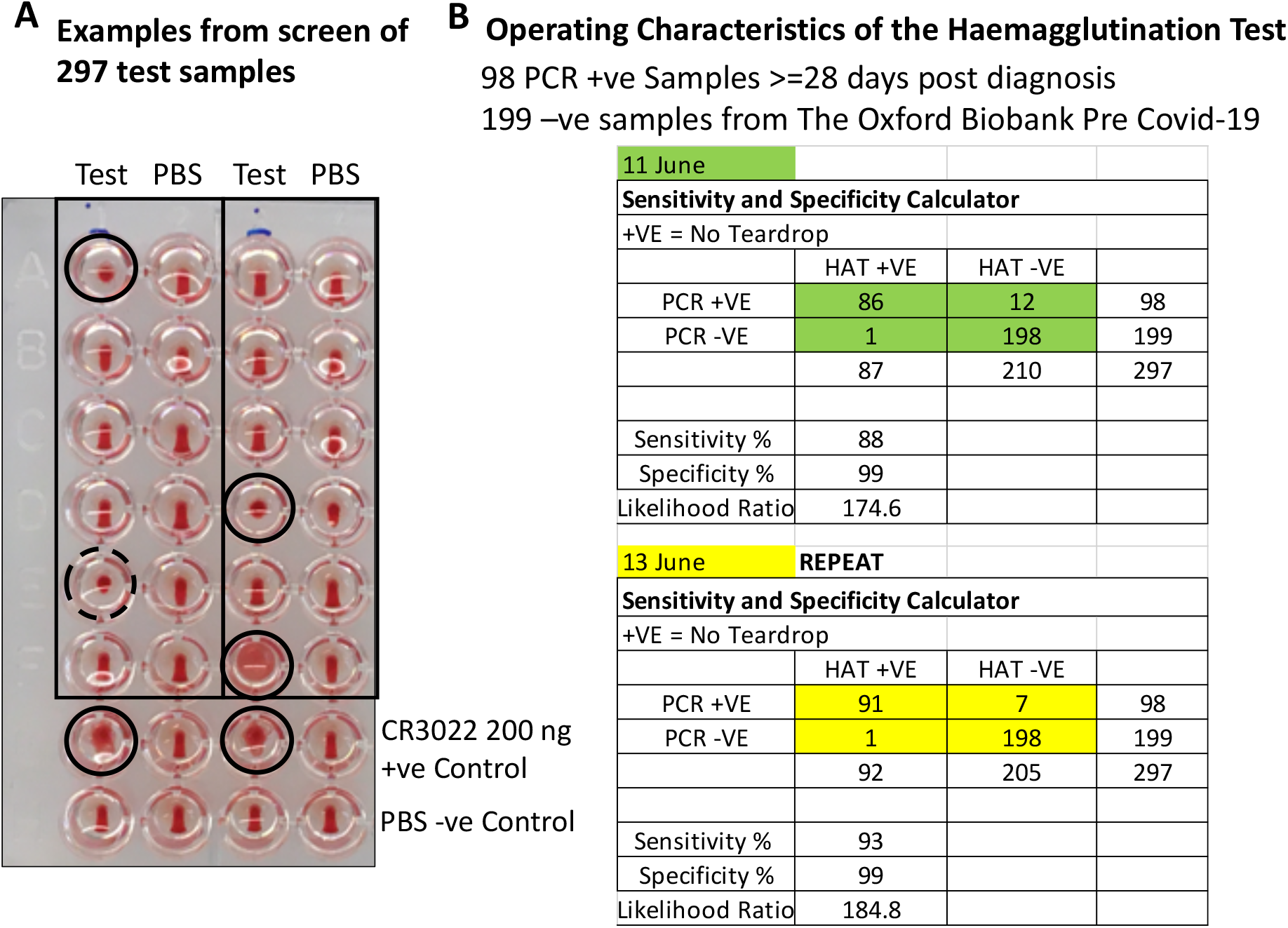
Operating characteristics of the HAT. A) The test set of 297 randomised plasma samples were diluted 1:40 mixed with 1:40 O-ve blood in two columns. IH4-RBD (100 ng in 50 μL) was added to the test samples, and PBS to negative control wells. The plates were incubated at room temperature for one hour to allow the red cell pellet to form, then tilted for ∼30 seconds to allow a teardrop to form. Complete loss of teardrop was scored as positive agglutination (marked with a black solid-line circle). Full teardrop or partial teardrop (marked with a dotted-line circle) were scored negative. The samples in columns were re-randomised and tested for a second time two days later. B) Contingency table showing the operating characteristics of the HAT.

With these rules in place we obtained in the first run sensitivity 88%, specificity 99%, and in the second run sensitivity 93%, specificity 99% (Figure 4). The Siemens Atellica Chemiluminescence assay for detection of IgG antibodies to the RBD was run in parallel on 293 of these 297 samples and gave sensitivity 100%, specificity 100% for this sample set.

We decided prior to this formal assessment to score wells with partial teardrop formation as negative, as these wells tended to give rise to disagreements between scorers and were not very helpful. Fifteen of 297 wells gave a partial teardrop. Six of these fifteen were from PCR-ve donors and scored negative on the Siemens assay, 9/16 were from PCR+ve donors and were Siemens positive. If partial teardrops were scored as positive, the sensitivity of the two assays increased to 97% and 99% (from 88% and 93%), but specificity was reduced to 96% and 98% (from 99%). This small loss of specificity would be unacceptable in sero-surveys where the expected prevalence of previous SARS-CoV-2 infection was low.

### HAT in the Hospital Setting

We next assessed the HAT in the setting of patients recently admitted to hospital (the first five days) through access to the COMBAT collection of samples (see methods). This set comprised 153 plasma samples from donors diagnosed with COVID-19 by PCR, with clinical syndromes classified as «Critical », «Severe », «Mild », and «PCR positive Health Care Workers ». Seventy-nine control plasma samples donated in the pre-COVID-19 era were obtained either from patients with bacterial sepsis (54 samples), or healthy volunteers (25 samples). Samples were titrated in 11 doubling dilutions of 50 μL from 1:40 – 1:40,096 (columns 1-11). Column 12 contained 50 μL PBS as a negative control. 50 μL of 1:40 O-ve whole blood was added, followed by 50 μL of 2 μg/mL IH4-RBD (100 ng/well). In parallel, all of the 153 samples from PCR positive donors were assessed by the Siemens Atellica Chemiluminescence test for antibodies to the RBD of the spike protein. The HAT scores (as the number of doubling dilutions of the sample required to reach the endpoint of complete loss of teardrop), and representative agglutination results are shown in figure 5A. In Figure 5B the HAT scores are plotted with their related Siemens test scores.

**Figure 5.**
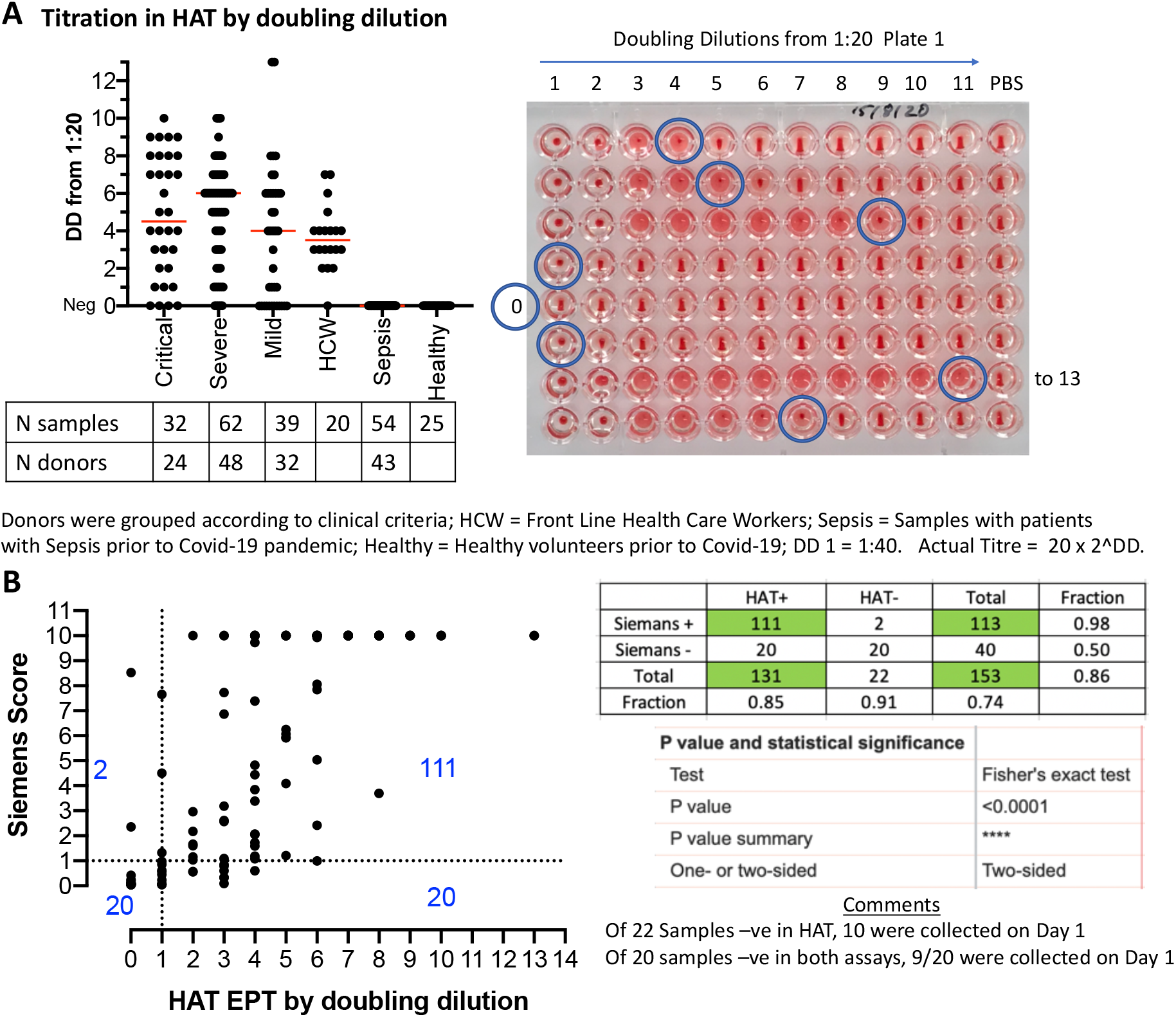
Titration of the set of 232 samples in the HAT. A) The collection included 32 samples from 24 Critical patients, 62 samples from 48 Severe, 39 samples from 32 Mild, 20 single samples from health care workers (HCW), 54 samples from 43 patients with unrelated sepsis in the pre-COVID-19 era, 25 samples from healthy unexposed controls. Median is indicated by a red line. DD: doubling dilutions. B) Comparison to Siemens Result (anti RBD) with HAT titre by doubling dilution for 153 samples from Critical, Severe, Mild and HCW SARS-CoV-2 PCR positive donors.

None of the seventy-nine negative control samples scored as positive in the HAT at a dilution of 1:40, thus providing 100% specificity in this set of samples. The HAT detected 131/153 (86% sensitivity) of the samples from PCR-diagnosed donors within the first five days of hospital admission, whereas the Siemens test detected 113/153 (74%). On day 5 the HAT detected 41/45 (91% sensitivity). Two samples had an endpoint greater than 11 doubling dilutions in the HAT, and required a repeat measurement spanning two plates. These two samples titrated to 13 doubling dilutions (1: 163,840). Unmasking the samples revealed that both were acquired from an elderly lady with mild disease on days 3 and 5 of her admission. The range of positive titres detected by HAT was broad: 1 to 13 doubling dilutions (1:40 – 1:163,840). A correlation coefficient with the Siemens test could not be calculated as the latter has a ceiling score of 10 (Figure 5B). A comparison of the two tests in a contingency table with cut-off of 1:40 (first doubling dilution) for HAT, and a score ≥ 1 for the Siemens test (as defined by the manufacturer), showed a strong correlation between the two tests for detection of antibodies to the RBD (P <0.0001; two-tailed Fisher’s exact test, Figure 5B). Fifty-two of the 153 samples were from 24 donors with COVID-19 from whom repeated samples were taken on days 1, 3, or 5 of admission. The HAT detected a rise in agglutination titre over the first five days of admission in 16/24 (67%) of these patients (Table 1). Reductions in titre were not detected.

**Table 1.** Fifty-two samples from 24 donors who were sampled repeatedly during the first five days in hospital.

|  |  | Day |  |  |  |  |
| --- | --- | --- | --- | --- | --- | --- |
| No | Case | 1 | 2 | 3 | 4 | 5 |
| 1 | C2 |  |  | 40 |  | 160 |
| 2 | C6 |  |  | 40 |  | 80 |
| 3 | C7 | 0 |  |  |  | 0 |
| 4 | C9 | 40 |  |  |  | 2560 |
| 5 | C10 | 320 |  | 320 |  |  |
| 6 | C23 | 0 |  | 160 |  | 640 |
| 7 | S5 | 80 |  | 160 |  |  |
| 8 | S6 | 1280 |  | 1280 |  | 2560 |
| 9 | S7 | 80 |  |  |  | 2560 |
| 10 | S12 | 640 |  | 1280 |  |  |
| 11 | S13 | 0 |  | 40 |  | 320 |
| 12 | S14 |  |  | 640 |  | 640 |
| 13 | S17 |  |  | 2560 |  | 2560 |
| 14 | S20 | 160 |  | 320 |  |  |
| 15 | S31 | 40 |  |  |  | 1280 |
| 16 | S41 | 640 |  | 1280 |  |  |
| 17 | S43 |  |  | 40 |  | 160 |
| 18 | S48 |  |  | 1280 |  | 2560 |
| 19 | M9 |  |  | 0 |  | 160 |
| 20 | M11 | 0 |  | 0 |  | 0 |
| 21 | M13 |  |  | 0 |  | 0 |
| 22 | M14 |  |  | 163840 |  | 163840 |
| 23 | M25 | 40 |  |  |  | 640 |
| 24 | M30 |  |  | 320 |  | 320 |

C: Critical, S: Severe, M: Mild

These results showed that in the setting of hospital admission in the UK for suspected COVID-19 disease, the HAT has an overall sensitivity of 86% and specificity of 100% by day five, and frequently (67%) detected a rise in HAT titre during the first five days of admission. In this context the HAT performed at least as well as the commercially available Siemens Atellica Chemiluminescence assay (74%) for the detection of antibodies to the RBD of SARS-CoV-2 spike protein. Twenty samples were negative in both tests, but nine of these were taken on day 1 of admission, which suggests that both tests have lower detection levels early in the course of hospital admission, before the antibody response has fully developed.

The O-ve blood used as indicator for this experiment was collected into a heparin tube, and then transferred to a K2EDTA tube. In order to be sure that the presence of heparin in the red cells had not altered the behaviour of the test, and to confirm the robustness of the results, we repeated the titrations on all of the 232 samples 34 days later, with fresh O-ve red cells from a different donor collected as usual into a K2EDTA tube. The results are shown in supplementary figures 2A-C. Specificity of the HAT remained at 100% (none of the 79 control samples were detected as positive at 1:40). The correlation with the previous assay was strong (R^2^ = 0.975), and 99% of the 232 titrations were within one doubling dilution of the matched earlier measurement. The slope of the correlation was 0.94 (95% CI 0.92-0.96), significantly less than 1. This was due to a proportion of results titrating to one doubling dilution lower titre. However, this had only a small impact on sensitivity (81% from 86%), which was still an improvement on the Siemens test (74%) in this context of the first five days of hospital admission. A rise in HAT titre in 16/24 during the first five days of admission was confirmed.

### HAT as a Point of Care Test on Capillary Samples

The HAT is designed to detect antibodies to the RBD starting at a serum dilution of 1:40, and we have found that that 50 μL of 1:40 dilution of whole blood provides an optimal concentration of red blood cells for detection by agglutination in V-bottomed 96 well plates. We have not completed an extensive analysis of the HAT as a Point of Care Test. However, we have preliminary evidence that lyophilised IH4-RBD sent to the National Institute of Immunology, New Delhi, functions as a Point of Care Test on capillary blood obtained by finger-prick. In Figure 6, three positive (donors 1, 2 and 3) and three negative (donors 4, 5 and 6) HAT results are compared to a standard ELISA for detection of antibodies to the RBD.

**Figure 6.**
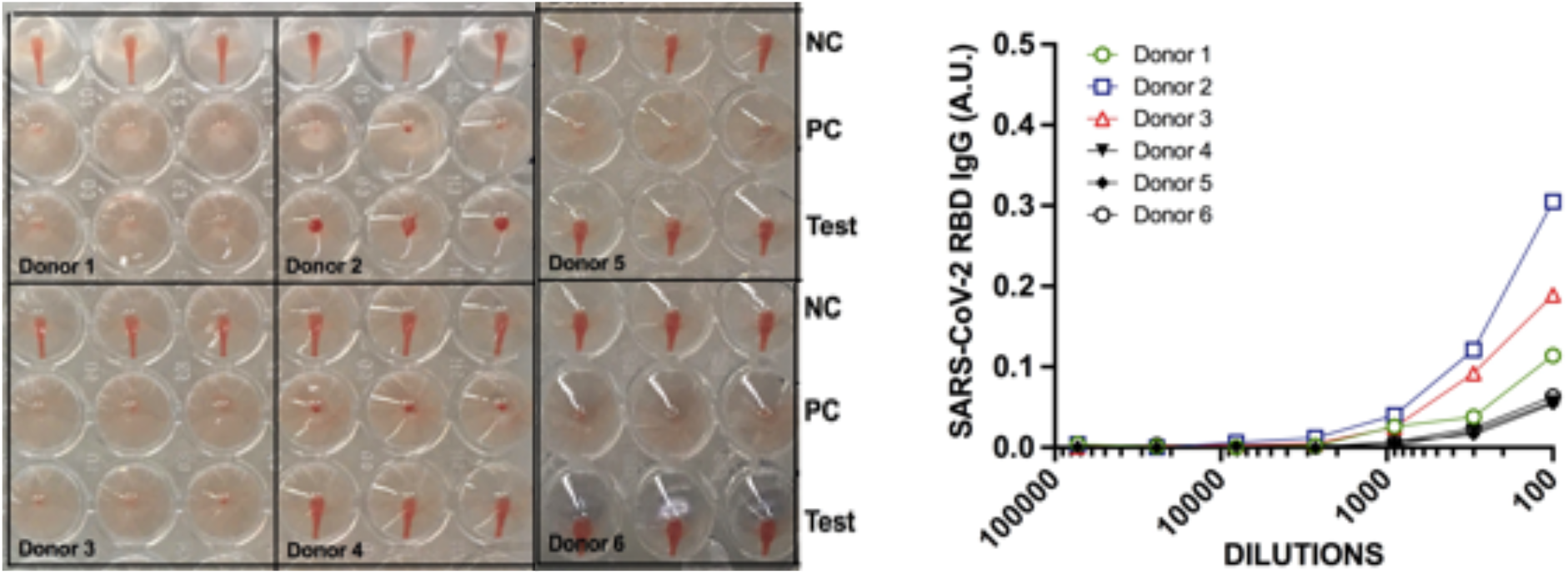
HAT as a point of care test. Capillary blood samples were obtained by lancet. Antibodies to the RBD were detected by HAT on autologous red cells in the sample in “Test” wells (plasma at 1:40) after addition of 100 ng/well IH4-RBD (see methods). NC, Negative Control (PBS replaces IH4-RBD); PC Postive Control (20 ng/well CR3022, an anti-RBD monoclonal antibody added). In parallel, after removal of red cells, the plasma was tested in a standard ELISA for detection of antibodies to the RBD. Low levels of antibody detected in the ELISA were sufficient to give a positive result in the HAT.

Further work is needed to establish the operating characteristics of the HAT as a Point of Care Test on capillary samples. We provide a suggested operating procedure for capillary samples in methods.

### Distribution of the IH4-RBD as lyophilised protein

In order to ship the IH4-RBD reagent efficiently we have examined the effects of lyophilisation and reconstitution with water. IH4-RBD synthesised for the purpose of distribution was provided at 5 mg/mL in PBS by Absolute Antibody Ltd, Oxford. Two hundred microlitre aliquots (1 mg, enough for 10,000 test wells) were lyophilised overnight and stored at −20 °C. Aliquots were thawed, reconstituted with 1 ml double distilled water and titrated against the pre-lyophilisation material. No change in the titration occurred. We have synthesised one gram of IH4-RBD, sufficient for 10 million test wells. This is available free of charge for any qualified group anywhere in the world in aliquots of 1 mg (20,000 test wells).

## DISCUSSION

The COVID-19 pandemic has had a particularly gruelling influence on the world economy, and on most populations of the world. The appearance of such a new highly contagious virus will probably not be a unique occurrence in the decades ahead. One of the lessons learned is the importance of developing affordable serological tests for detection of immune responses to SARS-CoV-2. Commercial antibody tests are not widely available to low- and middle-income countries, and lateral flow assays, while offering early promise as a near-patient test, have failed to deliver in terms of performance metrics, are expensive, and there are concerns about significant batch-to-batch variation (Adams et al., 2020). By contrast, the advantages of the HAT are the low cost of production of its single reagent (∼0.27 UK pence per test well), better performance than most lateral flow devices (Adams et al., 2020), and versatility in not requiring anything other than a source of O-ve blood (10 mL of K2EDTA blood provides enough for 8,000 tests), an adjustable pipette, PBS, and a standard 96 well V-bottomed plate.

We have demonstrated that the HAT functions as a viable test for the presence of antibodies to the RBD of the SARS-CoV-2 spike protein in stored serum/plasma samples, using O-ve red cells as indicators. In the formal assessment of sensitivity and specificity we recorded an average 90% sensitivity and 99% specificity (Likelihood Ratio ∼175), compared to 100% sensitivity and 100% specificity for the Siemens Atellica Chemiluminescence test on the same set of samples. The “positive” samples were selected to have been taken at least 28 days after a positive PCR test. These conditions are optimal for serological tests by allowing time for a rise in antibodies. The sensitivity of ∼90% and specificity of 99% did not reach the level of 98% for both recommended by the UK MHRA (The National SARS-CoV-2 Serology Assay Evaluation Group, 2020), or 99.5% for both recommended by the Infectious Disease Society of America (Hanson et al., 2020). However, these values are still consistent with a useful test in appropriate contexts, provided that users are fully aware of the operating characteristics and interpret the results correctly.

The sensitivity of the single point HAT can be enhanced (to ∼98%) if wells with partial teardrop formation are scored as positive. However, this improvement in sensitivity is gained at the expense of a reduced specificity (to ∼97%). If partial teardrops are to be scored as positive in the spot test at 1:40, we recommend obtaining confirmation for these by ELISA. Improvement in the operating characteristics of the HAT may be possible by a systematic analysis of buffer composition and experimental conditions, and is being investigated. We have deliberately kept complexity to a minimum, and thus all dilutions were made in standard PBS, and for the moment we recommend scoring wells with partial teardrop formation as negative.

It is interesting that the HAT titrations actually performed a little better than the Siemens test on 153 stored plasma samples from donors during the first five days of their hospital admission (note that symptom onset may have been several days earlier), in whom it detected 86% (81% in the repeat) of samples from PCR-diagnosed donors, compared to 74% for the Siemens test, and gave 100% specificity for the sample set containing control plasma from patients with sepsis and healthy controls. We speculate that at this early period of the COVID-19 illness the immune response may be dominated by IgM that would be expected to be particularly efficient at crosslinking the IH4-RBD labelled red cells. In addition, of the twenty-four donors who were tested more than once in the first five days in hospital, the HAT detected a rise in titre in sixteen (67%). A fixed high titre was detected in a further four to provide a sensitivity of 83% in these twenty-four cases. In situations of high clinical suspicion, the HAT could potentially have a place as a helpful test to support the diagnosis of COVID-19 by detecting a rising titre of antibodies to the RBD during hospital admission. In patients with a prior probability of a diagnosis of COVID-19 of ∼10%, the likelihood ratio of ∼175 for the HAT provides a posterior probability of ∼95% for this diagnosis. However, it is essential that if clinicians use a rising titre in the HAT as a diagnostic aid, they should be aware of the relatively low sensitivity (∼67%) in this context.

With a specificity of 99%, the HAT could be employed for epidemiological surveys of the seropositive rate in stored serum/plasma samples from populations with a moderate expected prevalence ∼10%, which would entail a negative predictive value of ∼90%. The HAT may be useful to detect seroconversion after vaccination, and for the identification of potential donors of high titre plasma for therapy, if the clinical trials that are in progress demonstrate a benefit. In the absence of knowledge about the level of antibody that indicates protection, the HAT should not be used to provide personal results to individuals, as discussed by the UK Royal College of Pathologists (https://www.rcpath.org/profession/on-the-agenda/COVID-19-testing-a-national-strategy.html). Finally, we show that the lyophilised IH4-RBD reagent sent to New Delhi functioned as expected in preliminary point of care testing on capillary samples obtained by finger-prick. However, additional evidence is needed to show that the sensitivity and specificity of the HAT, applied as a Point of Care Test in this way, are comparable to the tests on stored plasma samples, as stressed by the IDSA guideline on serological testing (Hanson et al., 2020). This will need to be done in field conditions, which is planned.

The technique required for applying the HAT can be learned in a day by a trained laboratory technician, paramedic, nurse or doctor. We have produced one gram of the developing IH4-RBD reagent (enough for ten million test wells) and offer to ship lyophilised aliquots of this material (sufficient for 10,000 tests) anywhere in the world, free of charge, for use as a research reagent for serological studies of COVID-19.

## METHODS

### Sample Collection and Ethics

**Figures 1,2:** Control whole blood (K2EDTA) as a source of red cells was collected from a healthy donor after informed consent.

**Figure 3:** Pre-pandemic negative controls: these samples were collected from healthy adults in the Oxfordshire region of the UK between 2014 and 2016, ethics approval: Oxfordshire Clinical Research Ethics Committee 08/H0606/107+5. Positive sample set: these were convalescent plasma donors recruited by NHS Blood and Transplant (NHSBT), ethics approval (NHSBT; RECOVERY [Cambridge East REC (ref: 20/EE/0101)] and REMAP-CAP [EudraCT 2015-002340-14] studies).

**Figures 4, 5 and Table 1:** Known COVID-19 positive samples were collated from three ethically approved studies: Gastro-intestinal illness in Oxford: COVID substudy [Sheffield REC, reference: 16/YH/0247]ISARIC/WHO, Clinical Characterisation Protocol for Severe Emerging Infections [Oxford REC C, reference 13/SC/0149], the Sepsis Immunomics project [Oxford REC C, reference:19/SC/0296]) and by the Scotland A Research Ethics Committee (Ref: 20/SS/0028). Patients were recruited from the John Radcliffe Hospital in Oxford, UK, between March and May 2020 by identification of patients hospitalised during the SARS-COV-2 pandemic and recruited into the Sepsis Immunomics and ISARIC Clinical Characterisation Protocols. Time between onset of symptoms and sampling were known for all patients and if labelled as convalescent patients were sampled at least 28 days from the start of their symptoms. Written informed consent was obtained from all patients. All patients were confirmed to have a test positive for SARS-CoV-2 using reverse transcriptase polymerase chain reaction (RT-PCR) from an upper respiratory tract (nose/throat) swab tested in accredited laboratories. The degree of severity was identified as mild, severe or critical infection according to recommendations from the World Health Organisation. Severe infection was defined as COVID-19 confirmed patients with one of the following conditions: respiratory distress with RR>30/min; blood oxygen saturation<93%; arterial oxygen partial pressure (PaO2) / fraction of inspired O2 (FiO2) <300 mmHg; and critical infection was defined as respiratory failure requiring mechanical ventilation or shock; or other organ failures requiring admission to ICU. Comparator samples from healthcare workers with confirmed SARS-CoV-2 infection who all had mild non-hospitalised disease were collected under the Gastro-intestinal illness in Oxford: COVID sub-study, and samples from patients with equivalently severe disease from non-COVID infection were available from the Sepsis Immunomics study where patients presenting with significantly abnormal physiological markers in the pre-pandemic timeframe had samples collected using the same methodology as that applied during the COVID pandemic. Blood samples were collected in K2EDTA vacutainers and PBMCs were separated from plasma using Sepmate isolation tubes (STEMCELL Technologies) and plasma was used in the downstream HAT assay.

**Figure 6:** Capillary samples were collected from members of institute staff with informed consent (New Delhi, India). The study is a part of the COVID-19 project ‘IPA/2020/000077’. The project has been approved by the Institutional Human Ethics Committee; Ref. no. - IHEC#128/20.

### Cloning, Expression and Purification of VHH(IH4)-RBD

The codon-optimised gene encoding IH4-RBD sequence (Figure 1B and supplementary for the cDNA sequence) was synthesized by Integrated DNA Technologies. The gene was cloned in to the AbVec plasmid (Genbank FJ475055) using the restriction sites AgeI and HindIII (the vector supplied the signal sequence). **This expression plasmid for IH4-RBD is available on request**. Protein was expressed in Expi293F™ cells using the manufacturer’s protocol (Thermo Fisher). Protein supernatant was harvested on day 5/6 after transfection, spun and 0.22 μm filtered. The protein was affinity-purified using a His-Trap HP column (Cytiva). Binding buffer consisted of 20 mM Sodium Phosphate, 150 mM NaCl and 20 mM Imidazole at pH 7.4, and the elution buffer of 500 mM Imidazole in 1 x binding buffer. Protein was concentrated using 15 ml Vivaspin 30 kDa MWCO filter and then buffer exchanged to PBS using a 10 ml 7 kDa Zebaspin column (Thermo Fisher).

For large scale production, the protein was synthesized by Absolute Antibody Ltd, Oxford using the same plasmid construct in HEK293 cells.

### Lyophilisation of IH4-RBD and CR3022 monoclonal antibody

For lyophilisation, 200 μL (1 mg) of IH4-RBD (5 mg/mL) and 100 μL (200 μg) CR3022 mAb (2 mg/mL) in PBS buffer prepared in Protein Lo-Bind microcentrifuge tube (Fisher Scientific) were frozen at −80 °C and further cooled down to - 196 °C using liquid nitrogen. Pre-cooled samples were transferred to Benchtop K freeze-dryer (VirTis) with chamber at 49 μbar and condenser pre-cooled to −72.5 °C. The samples were freeze-dried for a minimum of 24 h, wrapped in Parafilm (Merck) and stored at −20 °C. Lyophilised sample was reconstituted in the same original volume of MilliQ water.

### Indirect ELISA to detect SARS-CoV-2 specific IgG (Figure 6)

A standard indirect ELISA was used to determine the SARS-CoV-2 specific IgG levels in plasma samples. A highly purified RBD protein from SARS-CoV-2 Wuhan strain (NR-52306, BEI Resources, USA), expressed in mammalian cells, was used to capture IgG in the plasma samples. Briefly, ELISA plates (Nunc, Maxisorp) were coated with 100 μL/well of RBD antigen diluted in PBS (pH 7.4) at the final concentration of 1 μg/mL and incubated overnight at 4°C. Plates were washed three times with washing buffer (0.05% Tween-20 in PBS) followed by the incubation with blocking buffer (3% Skim milk and 0.05% Tween-20 in PBS). The 3-fold serially diluted heat inactivated plasma samples in dilution buffer (1% Skim milk and 0.05% Tween-20 in PBS) were added into the respective wells, followed by incubation at room temperature for 1 hour. After incubation, plates were washed, and anti-human IgG conjugated with Horseradish Peroxidase (HRP) (Southern Biotech) was added in each well. After 1 h incubation, plates were washed and developed by OPD-substrate (Sigma-Aldrich) in dark at room temperature. The reaction was stopped using 2N HCl and the optical density (OD) was measured at 492 nm. The RBD-antigen coated wells that were added with sample diluent alone were used as the blank. The OD values from sample wells were plotted after subtracting the mean of OD values obtained in the blank wells.

### Equipment and Reagents for HAT

- O-ve blood as a source of red cells collected in K2EDTA tube, diluted **in PBS to 1:20 or 1:40 as needed**. Resuspend by inverting gently ∼12 times.
- BD Contact Activated Lancet Cat. No. 366594 (2 mm x 1.5 mm)
- 100 μL, 20 μL pipettes, Multichannel pipettes
- V-bottomed 96-well plates (Greiner Bio-One, Cat. No. 651101, Microplate 96-well, PS, V-bottom, Clear, 10 pieces/bag)
- Eppendorf Tubes
- K2EDTA solution (add 5 mL PBS to 10 mL K2EDTA blood collection tube = 3.6 mg K2EDTA/mL, store at 4 °C)
- Phosphate Buffered Saline Tablets (OXOID Cat. No. BR0014G)
- IH4-RBD Reagent diluted 2 μg/mL in PBS. This remains active for at least 1-2 weeks stored at 4 °C
- V-bottomed 96-well plates, numbered, dated, timed (helps when timing many plates)
- Positive control monoclonal antibody CR3022 diluted to 2 μg/mL in PBS

### Other Reagents

Monoclonal antibody to human IgG (Gamma chain specific) Clone GG-5 Sigma Cat. No. I5885

#### 1. Spot test on Stored Serum/Plasma samples (Figure 4)

1. Plate out 50 μL of **1:20** serum/plasma in alternate columns 1,3,5,7,9,11 (add 2.5 μL sample to 47.5 μL PBS).
2. Add 50 μL **1:20** O-ve blood collected in (**so that now sample is diluted to 1:40 and red cells at ∼1% v/v**)
3. Mix and transfer 50/100 μL to neighbouring columns 2,4,6,8,10,12 for -ve controls. The negative control is important because in rare cases, particularly in donors who have received blood transfusions, the sample in principle may contain antibodies to non-ABO or Rhesus D antigens.
4. Add 50 μL IH4-RBD reagent (2 μg/mL in PBS = 100 ng/well) to Columns 1,3,5,7,9,11
5. Add 50 μL PBS to columns 2,4,6,8,10,12.
6. Inc 1 hr RT
7. Tilt for 30 seconds
8. Photograph: with mobile phone use the zoom function to obtain a complete field
9. Read as Positive = No teardrop, Negative <1:40 = partial teardrop, Neg = complete teardrop.
10. Two readers should read the plates independently, and disagreements resolved by taking the lesser reading.
11. For each batch of samples set up positive control wells containing 20-100 ng monoclonal antibody CR3022 (as in Finger-Prick test below). This establishes that all of the reagents are working.

#### 2. Titration of Stored Serum/Plasma Samples

1. Dilute samples to **1:20** in 50 μL PBS (2.5 μL to 47.5 μL) in V-bottomed plate in Rows A-H, column 1.
2. Prepare doubling dilutions with PBS across the plate columns 1-11 (1:40 to 1:40,960), PBS control in column 12. Eight samples can be titrated per 96-well plate.
3. Add 50 μL 1:40 O-ve red cells (1% v/v or 1:40 fresh EDTA O-ve blood sample) to all wells
4. Add 50 μL IH4-RBD (2 μg/mL, = 100 ng/well). [Note: the red cells and IH4-RBD can be pre-mixed and added together in either 50 μL or 100 μL volume, to save a step. This variation in technique does not alter the measured titres.]
5. Allow red cells to settle for 1 hr
6. Tilt plate for at least 30 s and photograph. The titre is defined by the last well in which the tear drop fails to form. Partial teardrop regarded as negative.

#### 3. Finger-prick test on capillary blood as a Point of Care Test

1. Preparation: Clean Hands, warm digit. Prepare a plate (96-well V-bottomed) labelled with Date and Time.
2. Prick skin on outer finger pulp with disposable, single use BD or another Lancet.
3. Wipe away first drop of blood with sterile towel/swab
4. Massage second drop
5. Take a minimum of 5 μL blood with 20 μL pipette, mix immediately into 20 μL K2EDTA (3.6 mg/mL/PBS) in Eppendorf. If possible, take 20 μL of blood and mix into 80 μL K2EDTA solution. Another approach is collection of blood drops into a BD Microtainer K2E EDTA lavender vials that take 250-500 μL.
6. For 5 μL sample dilute to 200 μL with PBS (add 175 μL PBS), for 20 μL sample dilute to 800 μL (add 700 μL PBS). ***Sample is now at 1:40, and the red cells are at the correct density (∼1% v/v assuming a haematocrit of 40%) to give a clear tear drop***.
7. Plate 50 μL x 3 in V bottomed microtitre wells labelled T (Test), + (PC, positive Control), - (NC, negative control) – see figure 6.
8. Add 10 μL of control anti RBD Mab CR3022 (2 μg/mL stock in PBS, 20 ng/well) to “+” well
9. Add 50 μL IH4-RBD (2 μg/mL in PBS) to “T” (Test) and “+ve” wells, 50 μL PBS to “-ve” well.
10. Incubate 1 hour at RT for Red Cells to form a pellet in the “-ve” well
11. Tilt plate against a well-lit white background for ∼30 seconds to allow Tear drop to form in “-ve” well.
12. The presence of antibodies to RBD is shown by loss of Tear Drop formation in the “T” and “+ve” wells. Occasionally a partial tear drop forms – these wells are counted as Negative.
13. **Photograph the plate to record the results with the date and time**. Results can be reviewed and tabulated later. Taking picture from a distance and using the zoom function helps to take a clear picture of all wells in a 96-well plate.
14. The negative (PBS) control should be done on every sample for comparison. The Positive control induced by CR3022 is used to check that all the reagents are working, and that the glycophorin epitope recognised by VHH(IH4) is present on the red cells. Absence of the IH4 epitope should be *very rare* (Habib et al., 2013). For setting up cohorts a positive control on every sample is therefore not necessary but should be included in every *batch* of samples.
15. If a 20 μL sample of blood was taken from the finger prick there should be 650 μL of the 1:40 diluted blood left. The red cells can be removed and a preparation of 1:40 O-ve red cells used as above to titrate the sample. In principle the autologous red cells could be washed repeatedly, resuspended in the same volume of PBS, and used as indicators for the titration, however we have not attempted to do this. The s/n is 1:40 plasma that can be used in confirmatory ELISA or other tests.

## Data Availability

All the data and reagents are available on request.

## Author Contributions

**Conceived, initiated and followed the project, documented portability and robustness of HAT method in a separate laboratory, recruited collaborators:** Etienne Joly.

**Designed and produced the IH4-RBD reagent, established conditions for lyophilisation, isolated and expanded human monoclonal antibodies to the RBD and ACE2-Fc, performed the standard HAT assays, wrote the paper:** Alain Townsend, Pramila Rijal, Julie Xiao, Tiong Kit Tan, Lisa Schimanski, Jiangdong Huo, Rolle Rahikainen, Kuan-Ying A Huang.

**Contributed examples of HAT as a Point of Care Test:** Nimesh Gupta.

**Provision of Serum/Plasma Sample Sets for Figure 4: Project management for Oxford serology work for sensitivity and specificity measurement, assessment of preliminary data:** Philippa Matthews, Derrick Crook, Sarah Hoosdally, Nicole Stoesser; **collection and processing of samples, coordination of sample banks and running Siemens assay, ethics, storage of pre-pandemic samples, coordination of provision of pre-pandemic samples from Oxford BioBank, and sero-positive donors through NHSBT :** Teresa Street, Justine Rudkin, Fredrik Karpe, Matthew Neville, Rutger Ploeg, David J Roberts, Abbie Bown, Richard Vipond, Marta Oliveira, Abigail A Lamikanra, Hoi Pat Tsang.

**Provision of Serum Sample Sets for Figure 5 and Table 1 (COMBAT samples):** Alexander J Mentzer, Julian C Knight, Andrew Kwok, Paul Klenerman, Christina Dold; **ISARIC4C Investigators:** J. Kenneth Baillie, Shona C Moore, Peter JM Openshaw, Malcolm G Semple, Lance CW Turtle; **Oxford Immunology Network Covid-19 Response Clinical Sample Collection Consortium:** Mark Ainsworth, Alice Allcock, Sally Beer, Sagida Bibi, Elizabeth Clutterbuck, Alexis Espinosa, Maria Mendoza, Dominique Georgiou, Teresa Lockett, Jose Martinez, Elena Perez, Veronica Sanchez, Giuseppe Scozzafava, Alberto Sobrinodiaz, Hannah Thraves.

## ACKNOWLEDGEMENT AND FUNDING

A.T. is funded by the Medical Research Council (MR/P021336/1), Townsend-Jeantet Charitable Trust (charity number 1011770) and the Chinese Academy of Medical Sciences (CAMS) Innovation Fund for Medical Science (CIFMS), China (grant no. 2018-I2M-2-002). N.G. is funded by the Science Engineering Research Board, Department of Science and Technology, India. P.C.M. is funded by the Wellcome Trust (grant ref 110110Z/15/Z). D.R. is supported by NIHR Oxford Biomedical Research Centre. National Institute for Health Research Biomedical Research Centre Funding Scheme (to G.R.S.), the Chinese Academy of Medical Sciences (CAMS) Innovation Fund for Medical Science (CIFMS), China (grant number: 2018-I2M-2-002). G.R.S. is supported as a Wellcome Trust Senior Investigator (grant 095541/A/11/Z).

The Spike Glycoprotein Receptor Binding Domain (RBD) from SARS-Related Coronavirus 2 Wuhan-Hu-1 (Figure 6), Recombinant from HEK293 Cells, NR-52306 was produced under HHSN272201400008C and obtained through BEI Resources, NIAID, NIH.

We thank the healthy volunteers who kindly donated their O-ve red cells for titration of samples.

## DECLARATION

Competing Interests none.

The views expressed are those of the author(s) and not necessarily those of the NHS, the NIHR, the Department of Health or Public Health England’.

## SUPPLEMENTARY DATA

**Supplementary Table 1.** Fifteen human monoclonal antibodies or nanobodies with endpoint titres for detection in the HA Test. One divalent nanobody VHH72-Fc and two monoclonal antibodies FD-5D and EZ-7A specific for the RBD failed to agglutinate red cells with the IH4-RBD reagent. Three monoclonal antibodies to other regions of the spike protein EW-8B, EW-9C and FJ-1C acted as negative controls.

| Mab or divalent<br>Nanobody | HAT endpoint titre<br>(ng/well) | Reference |
| --- | --- | --- |
| CR3022 | 2 | (Huo, Zhao, et al., 2020; ter Meulen et al., 2006) |
| EY-6A | 16 | (Zhou et al., 2020) |
| VHH-72-Fc | Negative | (Wrapp et al., 2020) |
| FI-4A | 16 | (Huang et al., 2020) |
| ACE2-Fc | 4 | (Huo, Le Bas, et al., 2020) |
| FI-3A | 8 | (Huang et al., 2020) |
| FI-1C | 4 | (Huang et al., 2020) |
| H11-H4-Fc | 16 | (Huo, Le Bas, et al., 2020) |
| FD-5D | Negative | (Huang et al., 2020) |
| FD-11A | 31 | (Huang et al., 2020) |
| FN-12A | 31 | (Huang et al., 2020) |
| FJ-10B | 62 | (Huang et al., 2020) |
| FM-7B | 62 | (Huang et al., 2020) |
| EZ-7A | Negative | (Huang et al., 2020) |
| S309 | 125 | (Pinto et al., 2020) |
| EW-8B non-RBD anti<br>Spike | Neg Control | (Huang et al., 2020) |
| EW-9C non-RBD anti<br>Spike | Neg Control | (Huang et al., 2020) |
| FJ-1C anti NTD | Neg Control | (Huang et al., 2020) |

**Supplementary Figure 1.**
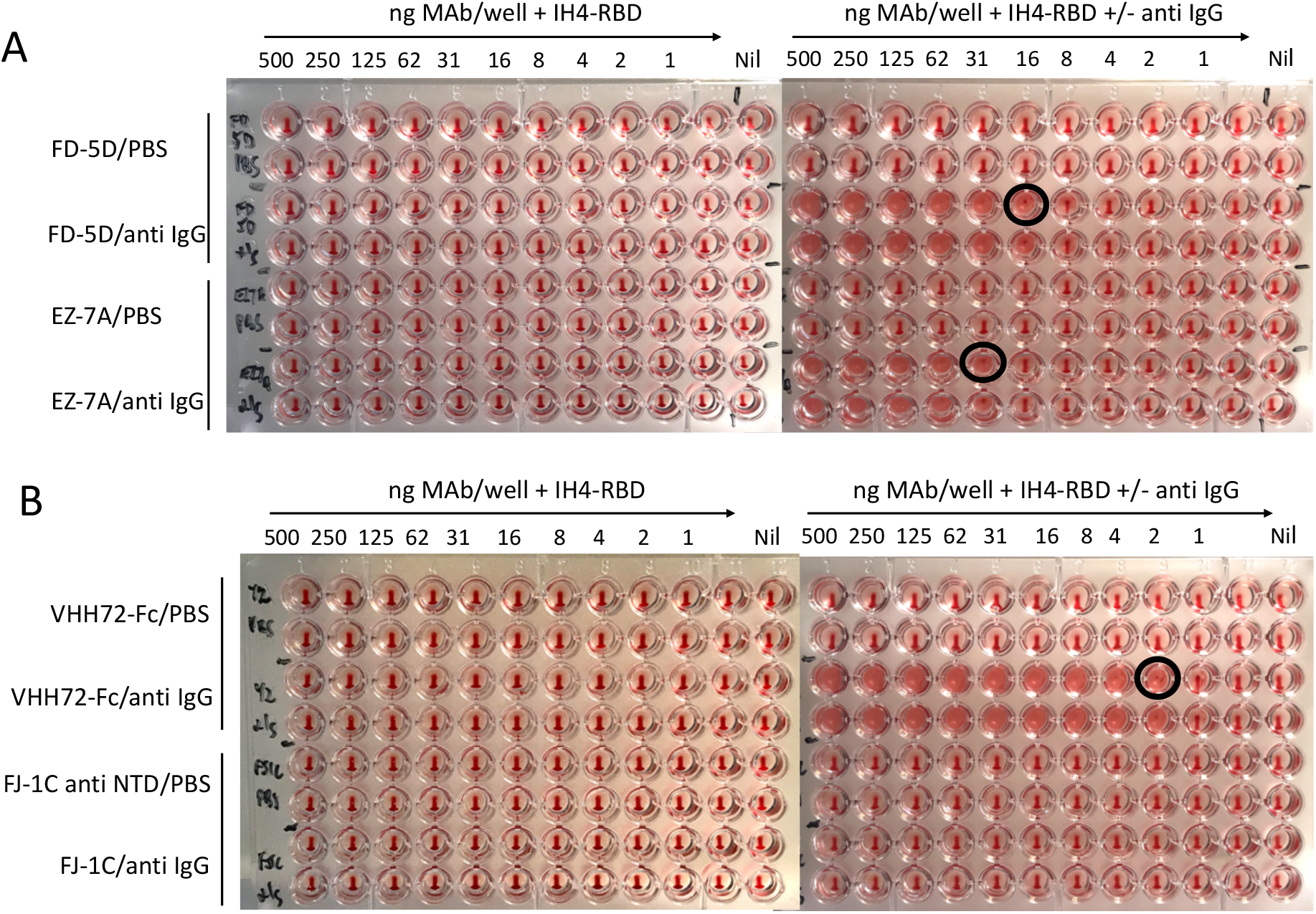
A, B) Three MAbs or Nanobody-Fc reagents of 15 tested specific for RBD failed to cross-link red cells but could be developed with an anti IgG reagent. Antibodies were titrated from 500 ng/well to 1 ng/well in 50 μL, 1:40 red cells were added in 50 μL, followed by 100 ng/well IH4-RBD in 50 μL, and incubated for 1 hour. The labelled red cells were then resuspended, and either 50 μL PBS (L hand plates), or Sigma Clone GG5 anti human IgG 1:100 in 50 μL (R hand plates). Red cells were allowed to settle for one hour, the plate tilted for 30s and photographed.

**Supplementary Figure 2.**
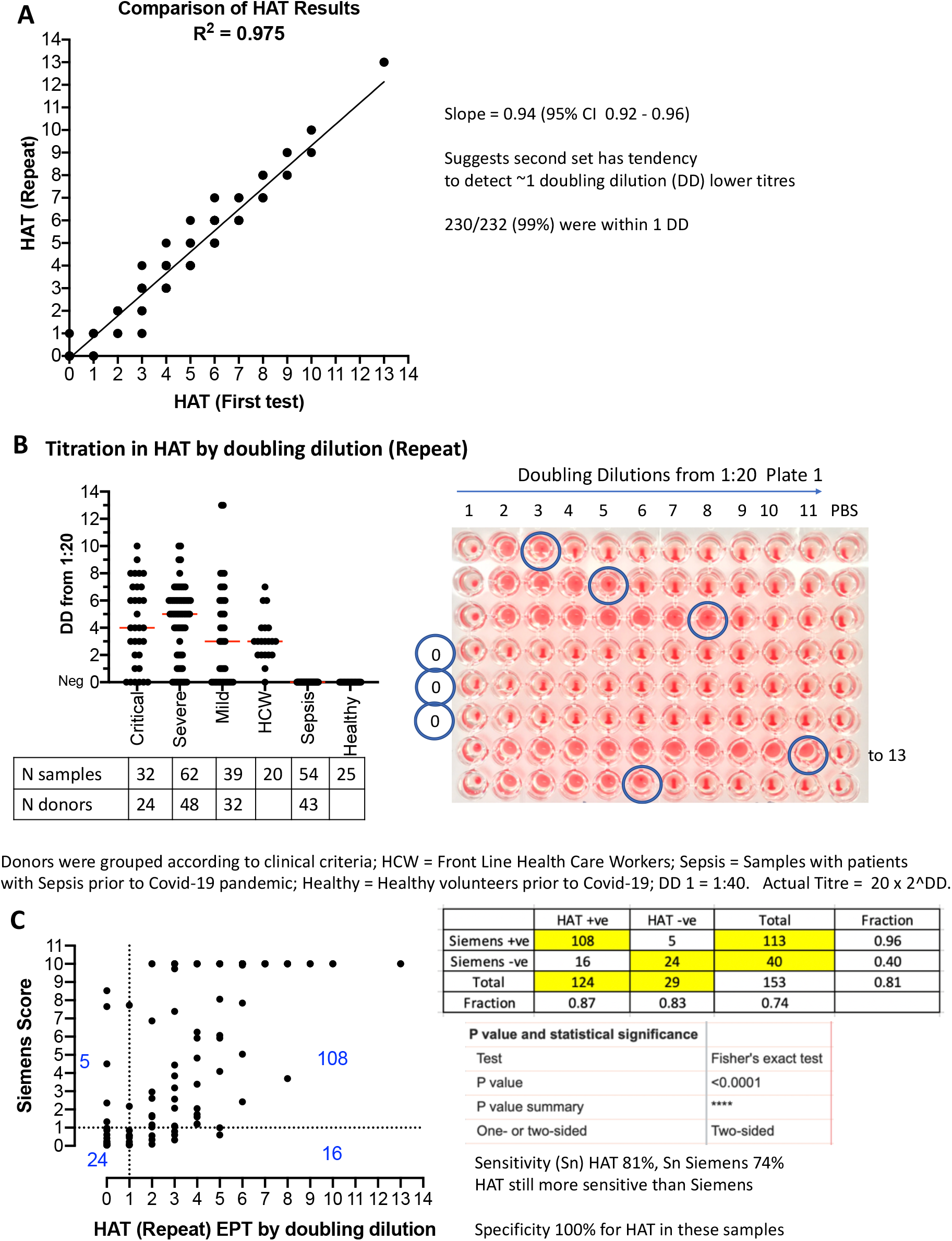
A) The 232 samples from figure 5 in the main paper were titrated a second time 34 days after the first measurement, as described for figure 4 (Methods). The values for the end point doubling dilution were compared and a correlation coefficient, and slope calculated (Prism v8). B) The collection included 32 samples from 24 Critical patients, 62 samples from 48 Severe, 39 samples from 32 Mild, 20 single samples from health care workers (HCW), 54 samples from 43 patients with unrelated sepsis in the pre-Covid-19 era, 25 samples from healthy unexposed controls. Median is shown. C) Comparison to Siemens Result (anti RBD) with HAT Titre by Doubling Dilution for 153 samples from Critical, Severe, Mild and HCW PCR+ve donors. The sensitivity of the HAT in this repeat was 81%, v 86% in the first test.

**Supplementary Table 2.** Fifty-two samples from 24 donors who were sampled repeatedly during the first five days in hospital; assay repeated 34 days after the measurements in Table 1. Sixteen of 24 showed a rise in titre.

|  |  | Day |  |  |  |  |
| --- | --- | --- | --- | --- | --- | --- |
| No | Case | 1 | 2 | 3 | 4 | 5 |
| 1 | C2 |  |  | 40 |  | 160 |
| 2 | C6 |  |  | 0 |  | 80 |
| 3 | C7 | 0 |  |  |  | 0 |
| 4 | C9 | 0 |  |  |  | 1280 |
| 5 | C10 | 160 |  | 320 |  |  |
| 6 | C23 | 0 |  | 80 |  | 320 |
| 7 | S5 | 40 |  | 40 |  |  |
| 8 | S6 | 1280 |  | 2560 |  | 2560 |
| 9 | S7 | 80 |  |  |  | 1280 |
| 10 | S12 | 320 |  | 640 |  |  |
| 11 | S13 | 0 |  | 0 |  | 160 |
| 12 | S14 |  |  | 640 |  | 1280 |
| 13 | S17 |  |  | 1280 |  | 1280 |
| 14 | S20 | 80 |  | 320 |  |  |
| 15 | S31 | 0 |  |  |  | 640 |
| 16 | S41 | 640 |  | 1280 |  |  |
| 17 | S43 |  |  | 40 |  | 80 |
| 18 | S48 |  |  | 1280 |  | 1280 |
| 19 | M9 |  |  | 40 |  | 160 |
| 20 | M11 | 0 |  | 0 |  | 0 |
| 21 | M13 |  |  | 0 |  | 0 |
| 22 | M14 |  |  | 163840 |  | 163840 |
| 23 | M25 | 40 |  |  |  | 320 |
| 24 | M30 |  |  | 160 |  | 160 |

## DNA sequence of codon optimised cDNA encoding IH4-RBD

ATGGGATGGTCATGTATCATCCTTTTTCTAGTAGCAACTGCAACCGGTGTTCATAGCCAGGTCCAGCTGCAAGAGTCTGGCGGAGGATCTGTTCAGGCTGGCGGAAGCCTGAGACTGAGCTGTGTGGCCAGCGGCTACACCGATAGCACATACTGCGTCGGCTGGTTCAGACAGGCCCCTGGCAAAGAGAGAGAGGGCGTCGCCAGAATCAACACCATCAGCGGCAGACCTTGGTACGCCGACTCTGTGAAGGGCAGATTCACAATCAGCCAGGACAACAGCAAGAACACCGTGTTTCTGCAGATGAACAGCCTGAAGCCAGAGGACACCGCCATCTACTACTGCACCCTGACCACCGCCAACAGCAGAGGCTTTTGTTCCGGCGGCTACAACTACAAAGGCCAGGGCACCCAAGTGACCGTGTCTAGCGCGTCGACCGGCTCTGGCGGCAGCGGCAACATCACCAATCTGTGCCCTTTCGGCGAGGTGTTCAACGCCACCAGATTTGCCAGCGTGTACGCCTGGAACCGGAAGAGAATCAGCAACTGCGTGGCCGACTACAGCGTGCTGTACAATAGCGCCAGCTTCAGCACCTTCAAGTGCTACGGCGTGTCCCCTACCAAGCTGAACGACCTGTGCTTCACCAATGTGTACGCCGACAGCTTCGTGATCAGAGGCGACGAAGTTCGGCAGATCGCTCCTGGACAGACAGGCAAGATCGCCGATTACAACTACAAGCTGCCCGACGACTTCACCGGCTGCGTGATCGCCTGGAATAGCAACAACCTGGACAGCAAAGTCGGCGGCAACTACAACTACCTGTACCGGCTGTTCCGGAAGTCCAACCTGAAGCCTTTCGAGCGGGACATCAGCACCGAGATCTATCAGGCCGGCAGCACCCCTTGTAATGGCGTGGAAGGCTTCAACTGCTACTTCCCACTGCAGTCCTACGGCTTTCAGCCTACAAACGGCGTGGGCTACCAGCCTTATAGAGTGGTGGTGCTGAGCTTCGAACTGCTGCATGCCCCTGCTACCGTGTGCGGCCCTAAAAAACACC ATCACCACCACCATTGA

